# Sub-Optimal Oral Health, Multimorbidity and Access to Dental Care

**DOI:** 10.1101/2023.12.19.23300225

**Authors:** L Limo, K Nicholson, S Stranges, N Gomaa

## Abstract

**INTRODUCTION:** Emerging research on the links between sub-optimal oral health and multimorbidity (MM), or the co-existence of multiple chronic conditions, has raised queries on whether enhancing access to dental care may mitigate the MM burden, especially in older age. Here, we aim to assess the association between sub-optimal oral health and MM and whether access to dental care can mitigate the risk of MM in individuals with sub-optimal oral health.

**METHODS:** We conducted a cross-sectional analysis using data from the Canadian Longitudinal Study on Aging (CLSA) (n=44,815, 45-84 years old). Edentulism, self-reported oral health (SROH), and other oral health problems (e.g., toothache, bleeding gums), were each used as indicators of sub-optimal oral health. MM was defined according to the Public Health Agency of Canada as having 2 or more chronic conditions out of cancer, cardiovascular diseases, chronic respiratory diseases, diabetes, and mental illnesses. Variables for access to dental care included the number of dental visits within the last year, dental insurance status, and cost barriers to dental care. We constructed multivariable step-wise logistic regression models and interaction terms with 95% confidence intervals and estimated prevalence ratio (PR) to assess the associations of interest, adjusting for *a priori* determined sociodemographic and behavioural factors.

**RESULTS:** Each of the sub-optimal oral health indicators were significantly associated with MM (edentulism PR=1.48, 95%CI 1.31, 1.68; poor SROH PR=1.81, 95%CI 1.62, 2.01; other oral health problems PR = 1.91, 95%CI 1.78, 2.06). The magnitude of this association was exacerbated in individuals who lacked dental insurance, could not afford dental care, and those who reported fewer dental visits within the last year.

**CONCLUSION:** The association between sub-optimal oral health and MM may be exacerbated by the lack of access to dental care. Policies aiming to enhance access to dental care may help mitigate the risk of MM.

## INTRODUCTION

The oral-systemic health connection has increasingly come under the spotlight over the past decades due to its high relevance to health policy and clinical practice (Jin et al., 2016; Peres et al., 2019). Studies have consistently suggested that oral diseases, particularly periodontal diseases, are associated with a higher risk of several chronic conditions, including cardiovascular disease (Aldossri et al., 2021; Dietrich et al., 2017), diabetes (D’Aiuto et al., 2017; Oates et al., 2023), respiratory infections (Scannapieco & Cantos, 2016), and cognitive impairment (Rohani et al., 2023); although the evidence on causality remains arguable. A recent umbrella review on the evidence linking oral health conditions and systemic non-communicable diseases (NCD) concluded that individuals with oral diseases may be at an increased risk of NCDs (Botelho et al., 2022). More recently, there have been investigations into multimorbidity (MM), or the co-existence of two or more chronic conditions, and its association with oral diseases, with emerging studies suggesting the association of oral diseases with an increased risk of early MM development (Larvin et al., 2021). Studies on the connection between oral health and MM are becoming increasingly relevant given the higher risk of MM with aging, its major impact on quality of life, its link to an increased risk of mortality, and its significant direct and indirect costs to individuals and societies (Watt & Serban, 2020; Zhao et al., 2019). Studies on the association between oral health and MM in an aging population, however, remain scarce.

Several bidirectional pathways can link oral and chronic conditions such as diabetes and cardiovascular diseases, and by extension, MM. For example, pathophysiological mechanisms such as an increased oral and systemic inflammatory load and oxidative stress, may trigger or exacerbate the progression of oral diseases and/or chronic conditions (Gomaa et al., 2017). Other pathways may be psychological, potentially contributing to mental health conditions, where poor oral health and subsequently affected dental appearance has been shown to negatively impact self-esteem, quality of life, and social interactions (Moeller et al., 2015; Sabbah et al., 2018). Importantly, there are common risk factors to oral and related chronic conditions which lie within the social and living environments including poor diet, smoking, high sugar consumption and psychosocial stress (Gomaa, 2022; Sheiham & Watt, 2000), all of which concentrate in less affluent individuals, including underserved and marginalized populations (Fleming et al., 2023; Tsakos et al., 2023). These shared common risk factors point towards the need for innovative medical and dental integration models. The recently published World Health Organization (WHO) Global Oral Health Status Report underscores the imperative for universal access to oral health care as a pivotal strategy in addressing and mitigating oral and potentially non-oral health inequalities (WHO, 2022). Although evidence has significantly varied, access to dental care has particularly emerged as a potential intervention that may alter the risk for NCDs. While it has been argued that the systemic impact of the provision of dental care is unnecessarily overstated, enhancing access to dental care may extend beyond the oral cavity to help mitigate the risk of NCDs and MM, particularly in individuals with poor oral health (Watt & Serban, 2020).

To this end, our study aimed to assess the extent of the association between sub-optimal oral health and MM amongst middle-aged and older Canadians, and whether this relationship can be modified by indicators of access to dental care. Our hypothesis was that sub-optimal oral health would be significantly associated with MM, and that these associations would be exacerbated in those who report barriers to accessing dental care.

## METHODS

### Data source and study design

In this cross-sectional study, we sourced data from the Canadian Longitudinal Study on Aging (CLSA), a large, national, population-based cohort that aims to understand the biological, medical, psychological, social, lifestyle, and economic factors that influence healthy aging. CLSA includes a total of 51,338 women and men aged 45 to 85 years from all Canadian provinces, excluding Indigenous people living on First Nations’ reserves, institutionalized individuals, and full-time members of the Canadian military. For our study, we used data from the first-follow up cycle (2015 to 2018), from both CLSA cohorts; comprehensive (in-person interviews), and tracking (telephone interviews). Approval to analyze the data for the purposes of this project was obtained from the CLSA (Application Number: 2203002) and the Western University Health Science Research Ethics Board (HSREB) (Project Number: 120760).

### Variables

#### Sub-optimal oral health

We used 3 indicators from the CLSA oral health module to define sub-optimal oral health, including edentulism, self-reported oral health (SROH), and other OH problems. Edentulism was defined by answering ‘no’ to the question ‘Do you have one or more of your original teeth?’. SROH was defined by the question ‘In general, would you say that the health of your mouth is excellent, very good, good, fair, or poor?’. In agreement with the literature in dichotomizing this standardized measure, we categorized this into good and poor SROH (Bassim et al., 2020; Hakeberg & Wide Boman, 2017). The variable OH problems was defined as the experience of any of the following in the past 12 months: toothache, sore or bleeding gums, burning mouth, avoiding eating particular foods because of problems with mouth, or other problems with mouth, teeth or dentures.

#### Multimorbidity (MM)

We defined MM as having at least 2 of 5 chronic conditions including: cancer; cardiovascular diseases (heart diseases, stroke); chronic respiratory diseases (asthma, chronic obstructive pulmonary diseases); diabetes; and mental illnesses (mood, anxiety disorders) according to the latest public health definition of the Public Health Agency of Canada (Public Health Agency of Canada, 2019). We opted to use this definition to allow for the comparability of our findings to other studies on MM from the Canadian context (Wilk et al., 2021).

#### Access to dental care

This was indicated by 3 measures (i) dental care utilization; (ii) availability of dental insurance, and (iii) the affordability of dental care. Dental care utilization was defined using the CLSA question ‘When did you last visit a dental professional?’ We dichotomized this variable into those who had visited a dental professional within the last 12 months and those who have last visited a dental professional more than 12 months ago at the time of data collection. Similarly, we dichotomized the availability of dental insurance as ‘yes’ for respondents who reported having dental insurance, and ‘no’ for those who did not; and the affordability of dental care using the question ‘In the past 12 months, have you not gone to a dental professional because of the cost of care?’, for which the response options were either ‘yes’ or ‘no’.

#### Covariates

These included sociodemographic and behavioural variables that were determined *a priori*. Age was categorized into 4 age groups: 45-54, 55-64, 65-74, and 75+ years or older, while sex was categorized as males and females, as reported in the CLSA. Race/ethnicity was categorized as White and non-White, as there was only a small number of respondents in the other racial/ethnic groups to allow for meaningful interpretations. Other covariates included socioeconomic factors such as total household income per annum and level of education and indicators of health behaviours including smoking status and alcohol consumption.

### Statistical analysis

To make estimates generalizable to the Canadian population and to address the complexity of the survey design, we used trimmed and analytic weights for the descriptive analyses and multivariable analyses (CLSA, 2017). Our statistical analyses were carried out in sequential steps. First, we applied weighted descriptive statistics to calculate means and standard deviations for continuous variables and weighted percentages for categorical variables, stratified by sex. Next, multivariable step-wise logistic regression models were constructed to estimate prevalence ratios (PR) and 95% confidence intervals (CI) of the association between sub-optimal oral health and MM, adjusted for the aforementioned covariates. Then, to assess the modifying effect of indicators of access to oral health on the relationship between sub-optimal oral health and MM, we constructed interactions terms between sub-optimal oral health and each of the access to dental care indicators (Rothman, K. J., 2002; Vandenbroucke et al., 2007). All analyses were conducted using Stata v18.0 statistical software (StataCorp LLC., 2022).

## RESULTS

### Characteristics of study population

Weighted descriptive statistics are shown in Tables 1. Our sample (n=41,815) was composed mostly of females (51.36%) and White individuals (94.36%). In this sample of middle-aged and older adults, almost one third of participants were in the 55 to 64 years old age group (32.71%), followed by 65 to 74 years old age group (29.56%). The majority of participants had post-secondary education, and 89.97% were currently employed. Most participants reported being non-smokers (92.16%) and regular consumers of alcohol (56.37%). While most respondents had overall good oral health, almost 10% reported having poor SROH and/or being edentulous. However, the majority (62.25%) reported having had other oral health problems such as toothache, bleeding gums, or avoiding eating particular foods because of problems with mouth in the past 12 months. More than half of the participants (57.85%) had dental insurance coverage. However, out of those without dental insurance, a slightly higher proportion of females (42.68%) reported not having any dental insurance and avoiding dental care due to cost (12.9%) than males (40.09% and 9.98%, respectively). Meanwhile, MM affected 23.50% of this sample.

**Table 1.**
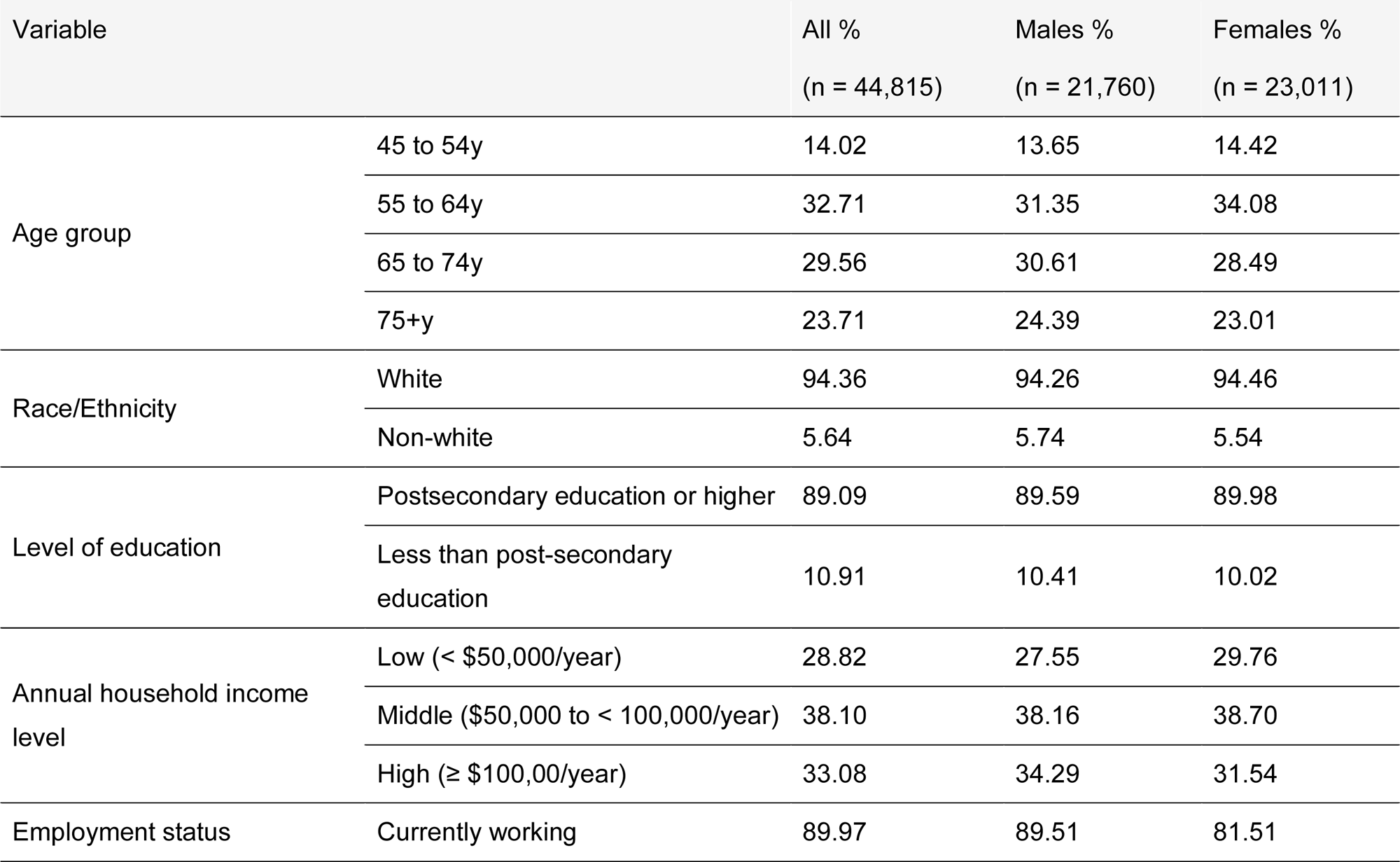
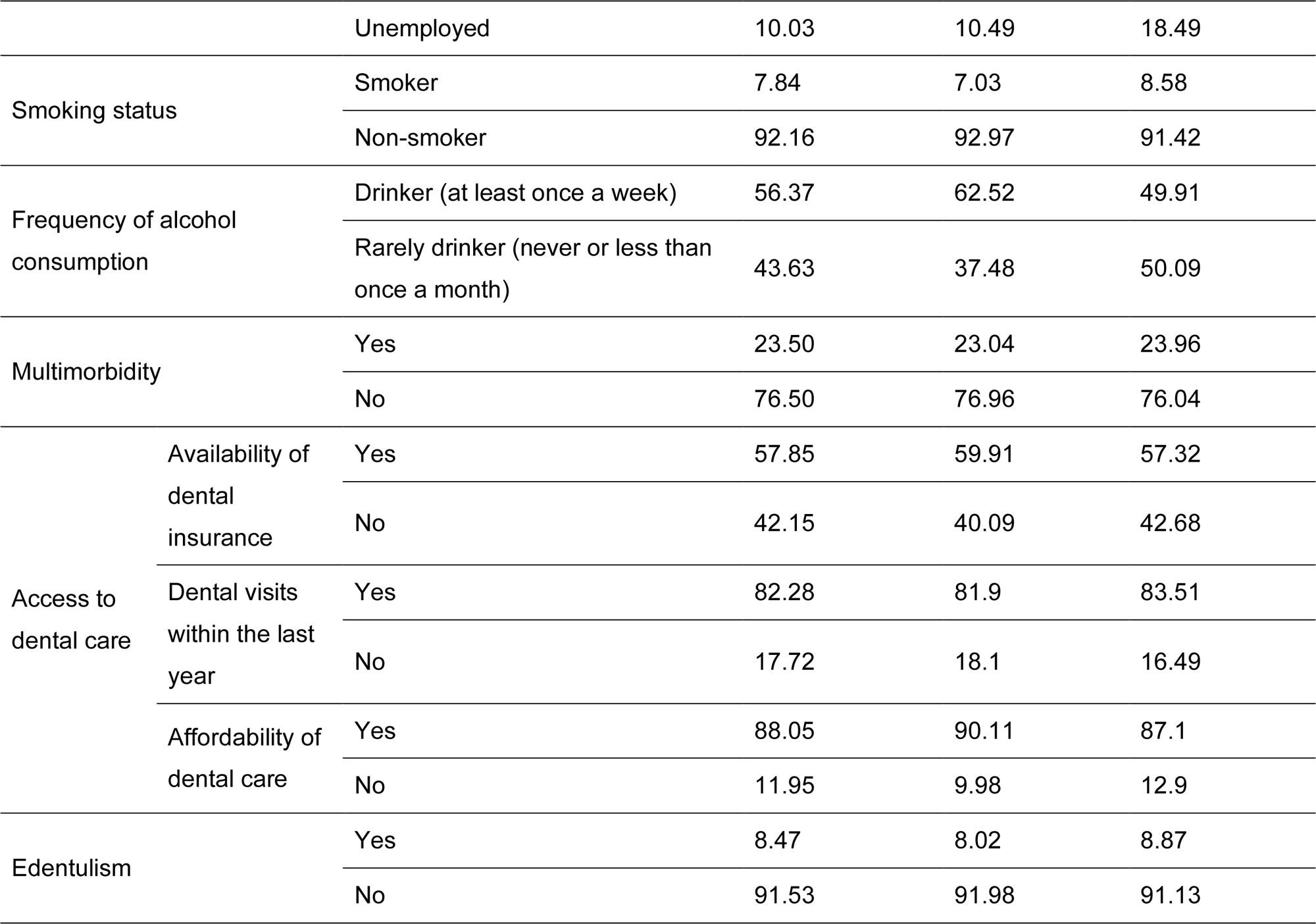
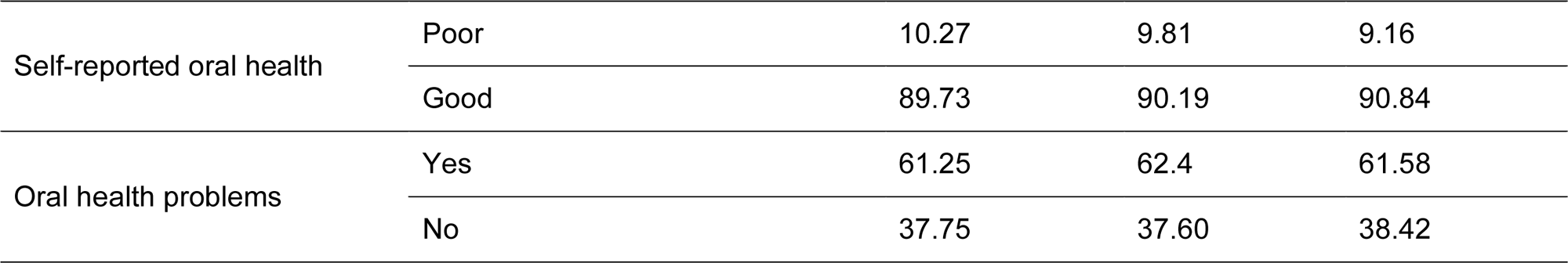
Weighted descriptive statistics of characteristics of study participants (n = 44,815), CLSA, first follow-up wave, 2018.

| Variable |  | All %<br>(n = 44,815) | Males %<br>(n = 21,760) | Females %<br>(n = 23,011) |
| --- | --- | --- | --- | --- |
| Age group | 45 to 54y | 14.02 | 13.65 | 14.42 |
|  | 55 to 64y | 32.71 | 31.35 | 34.08 |
|  | 65 to 74y | 29.56 | 30.61 | 28.49 |
|  | 75+y | 23.71 | 24.39 | 23.01 |
| Race/Ethnicity | White | 94.36 | 94.26 | 94.46 |
|  | Non-white | 5.64 | 5.74 | 5.54 |
| Level of education | Postsecondary education or higher | 89.09 | 89.59 | 89.98 |
|  | Less than post-secondary education | 10.91 | 10.41 | 10.02 |
| Annual household income level | Low (< \$50,000/year) | 28.82 | 27.55 | 29.76 |
| | Middle (\$50,000 to < 100,000/year) | 38.10 | 38.16 | 38.70 |
| | High (≥ \$100,00/year) | 33.08 | 34.29 | 31.54 |
| Employment status | Currently working | 89.97 | 89.51 | 81.51 |

|  |  |  |  |  |  |
| --- | --- | --- | --- | --- | --- |
|  |  | Unemployed | 10.03 | 10.49 | 18.49 |
| Smoking status |  | Smoker | 7.84 | 7.03 | 8.58 |
|  |  | Non-smoker | 92.16 | 92.97 | 91.42 |
| Frequency of alcohol consumption |  | Drinker (at least once a week) | 56.37 | 62.52 | 49.91 |
|  |  | Rarely drinker (never or less than once a month) | 43.63 | 37.48 | 50.09 |
| Multimorbidity |  | Yes | 23.50 | 23.04 | 23.96 |
|  |  | No | 76.50 | 76.96 | 76.04 |
| Access to dental care | Availability of dental insurance | Yes | 57.85 | 59.91 | 57.32 |
|  |  | No | 42.15 | 40.09 | 42.68 |
|  | Dental visits within the last year | Yes | 82.28 | 81.9 | 83.51 |
|  |  | No | 17.72 | 18.1 | 16.49 |
|  | Affordability of dental care | Yes | 88.05 | 90.11 | 87.1 |
|  |  | No | 11.95 | 9.98 | 12.9 |
| Edentulism |  | Yes | 8.47 | 8.02 | 8.87 |
|  |  | No | 91.53 | 91.98 | 91.13 |
| Self-reported oral health | Poor | 10.27 | 9.81 | 9.16 |
|  | Good | 89.73 | 90.19 | 90.84 |
| Oral health problems | Yes | 61.25 | 62.4 | 61.58 |
|  | No | 37.75 | 37.60 | 38.42 |

### Sub-optimal oral health increases the risk of MM

Each of the sub-optimal oral health outcomes was significantly associated with MM in unadjusted and fully adjusted models. Specifically, the crude estimates indicated almost twice the risk of MM in those reporting poor SROH (PR 2.01, 95% CI 1.82, 2.22), those experiencing other oral health problems in the past 12 months (PR 1.95, 95% CI 1.82, 2.09) and edentulous participants (PR 1.94, 95% CI 1.74, 2.17). These associations remained significant in models that were fully adjusted for socioeconomic factors, smoking and alcohol consumption (Table 2).

**Table 2.** Crude and adjusted association between indicators of sub-optimal oral health and MM. CLSA, first follow-up wave, 2018.

|  | PR<sub>1</sub> (95% CI) | PR<sub>2</sub> (95% CI) | PR<sub>3</sub> (95% CI) | PR<sub>4</sub> (95% CI) |
| --- | --- | --- | --- | --- |
| Edentulism | 1.94* (1.74, 2.17) | 1.64* (1.47, 1.83) | 1.44* (1.27, 1.62) | 1.40* (1.24, 1.58) |
| Poor SROH | 2.01* (1.82, 2.22) | 1.99* (1.81, 2.20) | 1.80* (1.61, 2.00) | 1.80* (1.61, 2.00) |
| Other oral health problems | 1.95* (1.82, 2.09) | 1.98* (1.85, 2.12) | 1.91* (1.77, 2.05) | 1.90* (1.76, 2.05) |

\* $p$ -value $\leq 0.05$
1: unadjusted (crude)
2: adjusted for age, sex, and race/ethnicity
3: adjusted for age, sex, race/ethnicity, income level, education level
4: adjusted for age, sex, race/ethnicity, income level, education level, smoking status, alcohol consumption

### Factors of access to dental care exacerbate MM in those with sub-optimal oral health

We generated interaction terms between each of the sub-optimal oral health indicators and the access to dental care variables to assess the impact of the latter on the risk of MM in fully adjusted models (Table 3). Analyses showed participants with poor SROH who had no dental insurance to be at a 1.54 times greater risk of MM than their counterparts with dental insurance. Similarly, participants with poor SROH who reported avoiding seeing the dentist due to costs were at a 1.63 higher risk of MM than those who reported no cost barriers to dental care. Meanwhile, edentulous participants who reported not being able to visit the dentist within the last year had 1.26 times higher risk of MM than their counterparts who had visited the dentist within the last year. Similar risk of MM was observed in participants who reported having other oral health problems over the past 12 months while having barriers to accessing dental care.

**Table 3.**
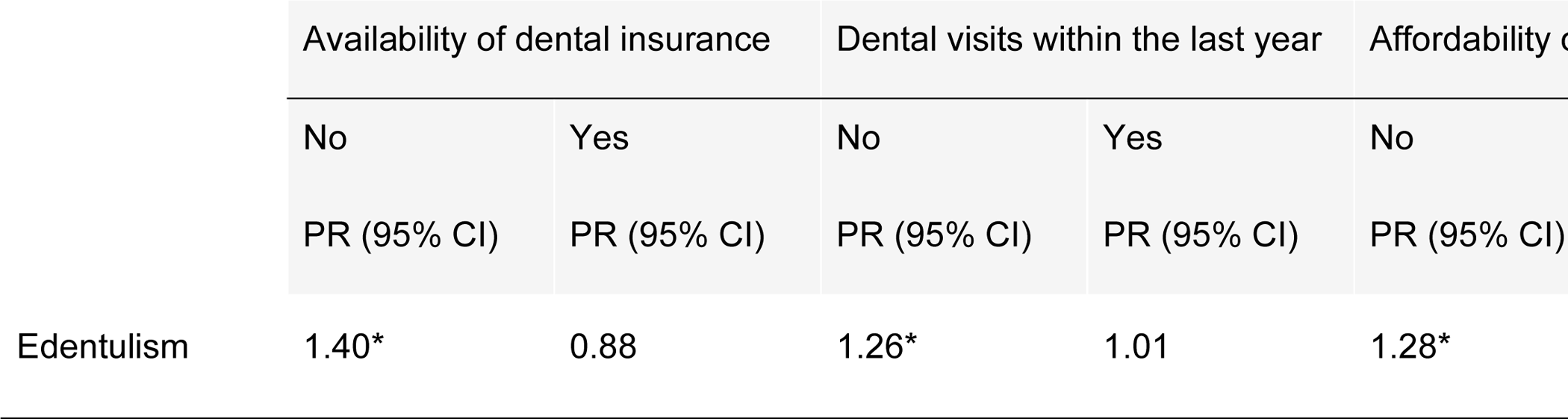
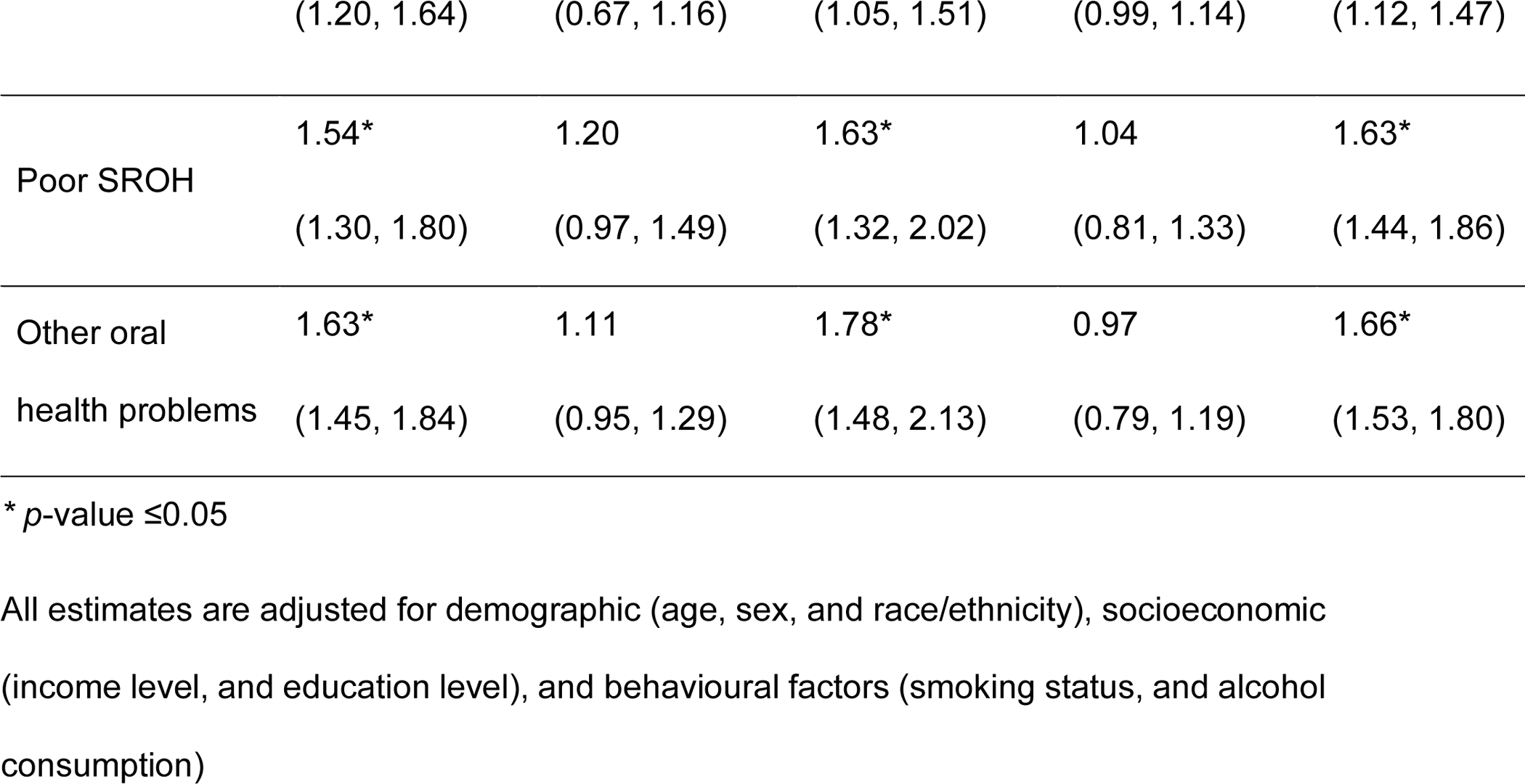
Modification of the association of sub-optimal oral health and MM by indicators of access to dental care. CLSA, first follow-up wave, 2018.

## DISCUSSION

In this study, we aimed to investigate the association between sub-optimal oral health and MM, and to assess the impact of factors of access to dental care on this association in a large, population-based sample of middle-aged and older Canadians. We conducted a cross-sectional analysis of data retrieved from the first follow-up wave of the CLSA, in which we found edentulism, poor SROH, and other oral health problems to have twice as high the risk of MM than their counterparts with overall good oral health outcomes. As hypothesized, we found the lack of access to dental care including having no dental insurance, not visiting a dentist within the last year, and avoiding dental care due to costs to exacerbate the risk of MM in respondents with sub-optimal oral health.

In agreement with previous research on oral health inequalities in Canada, we found issues of access to dental care such as financial barriers to dental care and to lack of dental insurance to be more common in females than males (Allison, 2023; Canadian Academy of Health Sciences, 2014). This may be accentuated by gender income inequalities and the greater barriers in accessing health care that affect females, due to factors of limited insurance coverage, caregiving responsibilities, and others (Manuel, 2018; Northridge et al., 2020; Shaw et al., 2017). These barriers may result in delayed or inadequate treatment, leading to increased disease burden (Statistics Canada, 2023).

Previous research has emphasized the impact that poor oral health and tooth loss can have on systemic health through impairing nutrition, digestion, and metabolism (Polzer et al., 2010; Zelig et al., 2022). Moreover, poor oral health and tooth loss affect the quality of life and well-being through the resultant discomfort, aesthetic problems, social isolation, and psychological distress (McKenna et al., 2020). Our results on the links between sub-optimal oral health and MM are in agreement with previous studies showing oral health conditions to be associated with the risk for other non-oral chronic conditions, such as cardiovascular disease, diabetes, respiratory infections, and mental illnesses (Felton, 2016; Watt & Aida, 2022). Importantly, studies on the oral-systemic health connection have been arguably controversial. While both groups of health conditions are linked, attributing systemic conditions to oral diseases has been consistently deemed as an overstatement, and that any connections are mostly due to the structural and social risk factors that are common to both conditions, including proper access to quality health care (Tsakos & Quiñonez, 2013). In that regard, our analyses suggest that enhancing access to dental care such as providing dental insurance and eliminating cost barriers to dental care may modify the sub-optimal oral health-MM relationship, or in other words, reduce the risk of MM in those with sub-optimal oral health.

Various mechanisms can underlie the role of dental care in reducing the risk of MM in individuals with sub-optimal oral health. Reducing inflammation, controlling infection, and promoting healthy behavioural changes can take place through enhanced access to dental care. While there is scarcity of studies on the impact of access to dental care on MM, our findings are supported by previous cross-sectional studies assessing the impact of dental care on chronic conditions. For example, a study using data from the National Health and Aging Trends Study (NHATS) from the United States found that dental care use was associated with lower odds of hypertension, diabetes, heart disease, stroke, and chronic obstructive pulmonary disease (COPD), adjusted for confounders such as age, sex, race, education, income, smoking, and body mass index (Meyerhoefer et al., 2021). While our results particularly focussed on the role of access to dental care in mitigating MM in individuals with sub-optimal oral health, it is also important to note that access to dental care may not be “the magic bullet” that can resolve chronic disease risk. A previous study in an elderly population in Japan, found the regular utilization of dental services to be associated with a higher number of remaining teeth in older age, however, not the presence of systemic conditions (Ohi et al., 2009). Surely, the risk of chronic conditions, and thereby MM, is multifactorial and dental attendance may be influenced by various factors related to oral health status, general health status, and socio-demographic characteristics (Peres et al., 2019; Watt et al., 2019). Hence, the upstream determinants of health should be regarded as key areas of investigation and intervention (Barnett et al., 2012; Elani et al., 2018).

The strengths of our study include our use of a large, population-based sample from a national survey i.e. the CLSA, as well as employing the most recent public health definition of MM as suggested by PHAC (Public Health Agency of Canada, 2019) thereby adding to the comparability of our results to previous studies on MM in Canada (Im et al., 2022; Nicholson et al., 2020). We also acknowledge several limitations, such as the cross-sectional study design due to the oral health data availability in CLSA which only allowed us to use the follow-up wave to address our research questions. Due to the lack of racial/ethnic diversity in CLSA, population groups including recent migrants and individuals from non-White racial/ethnic backgrounds may be underrepresented (Raina et al., 2019). In consequence, the results of our study cannot be generalized to these groups. It is also important to note that the oral health variables we used are not mutually exclusive, meaning that individuals who reported being edentulous may have also reported having poor SROH or having other oral health problems.

Furthermore, our study has implications for policy and practice. Our findings suggest that improving oral health and access to dental care for people who need it most may have beneficial effects through mitigating the risk of MM. Particularly, the links between sub-optimal oral health and MM are suggestive of that medical-dental integration to improve health outcomes for vulnerable populations such as those who lack access to dental care may be key. Policies and public health programs should address the social determinants of health and related health system factors such as dental insurance, that can contribute to oral and general health. With the latest WHO Global Oral Health Status Report calling for universal access to oral health care (WHO, 2022), and a new Canadian Federal dental care plan kicking-off, it will be important for future research to employ natural experiments using CLSA follow-up waves and methods of causal inference to further investigate the long-term impacts of improved access to dental care on MM.

## CONCLUSIONS

The association of sub-optimal oral health with MM in this population was exacerbated in individuals facing barriers to dental care. Enabling access to dental care may help mitigate MM in middle-aged and older Canadians with oral health problems.

## Data Availability

All data produced in the present work are contained in the manuscript

## ACKNOWLEDMENT

NG is supported by funding from the Schulich School of Medicine & Dentistry, Western University and the Lawson Health Research Institute, London, Ontario, Canada. This research was made possible using data collected by the Canadian Longitudinal Study on Aging (CLSA). Funding for the CLSA is provided by the Government of Canada through the Canadian Institutes of Health Research (CIHR) under grant reference: LSA 94473 and the Canada Foundation for Innovation, as well as the following provinces, Newfoundland, Nova Scotia, Quebec, Ontario, Manitoba, Alberta, and British Columbia. This research has been conducted using the CLSA first follow-up from the Comprehensive Dataset version 3.2 and Tracking Dataset version 2.3, under Application Number [2203002]. The CLSA is led by Drs. Parminder Raina, Christina Wolfson, and Susan Kirkland.

## REFERENCES

Aldossri, M., Farmer, J., Saarela, O., Rosella, L., & Quiñonez, C. (2021). Oral Health and Cardiovascular Disease: Mapping Clinical Heterogeneity and Methodological Gaps. JDR Clinical & Translational Research, 6(4), 390–401. 10.1177/2380084420953121

Allison, P. J. (2023). Canada’s oral health and dental care inequalities and the Canadian Dental Care Plan. Canadian Journal of Public Health. 10.17269/s41997-023-00800-6

Barnett, K., Mercer, S. W., Norbury, M., Watt, G., Wyke, S., & Guthrie, B. (2012). Epidemiology of multimorbidity and implications for health care, research, and medical education: A cross-sectional study. Lancet (London, England), 380(9836), 37–43. 10.1016/S0140-6736(12)60240-2

Bassim, C. W., MacEntee, M. I., Nazmul, S., Bedard, C., Liu, S., Ma, J., Griffith, L. E., & Raina, P. (2020). Self-reported oral health at baseline of the Canadian Longitudinal Study on Aging. Community Dentistry and Oral Epidemiology, 48(1), 72–80. 10.1111/cdoe.12506

Botelho, J., Mascarenhas, P., Viana, J., Proença, L., Orlandi, M., Leira, Y., Chambrone, L., Mendes, J. J., & Machado, V. (2022). An umbrella review of the evidence linking oral health and systemic noncommunicable diseases. Nature Communications, 13(1), Article 1. 10.1038/s41467-022-35337-8

Canadian Academy of Health Sciences. (2014). Improving access to oral health care for vulnerable people living in Canada – Canadian Academy of Health Sciences | Académie canadienne des sciences de la santé. https://cahs-acss.ca/improving-access-to-oral-health-care-for-vulnerable-people-living-in-canada/

Canadian Longitudinal Study on Aging. (2017). Sampling and Computation of Response Rates and Sample Weights for the Tracking (Telephone Interview) Participants and Comprehensive Participants. https://www.clsa-elcv.ca/doc/1041

D’Aiuto, F., Gable, D., Syed, Z., Allen, Y., Wanyonyi, K. L., White, S., & Gallagher, J. E. (2017). Evidence summary: The relationship between oral diseases and diabetes. British Dental Journal, 222(12), Article 12. 10.1038/sj.bdj.2017.544

Dietrich, T., Webb, I., Stenhouse, L., Pattni, A., Ready, D., Wanyonyi, K. L., White, S., & Gallagher, J. E. (2017). Evidence summary: The relationship between oral and cardiovascular disease. British Dental Journal, 222(5), Article 5. 10.1038/sj.bdj.2017.224

Elani, H. W., Simon, L., Ticku, S., Bain, P. A., Barrow, J., & Riedy, C. A. (2018). Does providing dental services reduce overall health care costs?: A systematic review of the literature. The Journal of the American Dental Association, 149(8), 696–703.e2. 10.1016/j.adaj.2018.03.023

Felton, D. A. (2016). Complete Edentulism and Comorbid Diseases: An Update. Journal of Prosthodontics, 25(1), 5–20. 10.1111/jopr.12350

Fleming, E., Bastos, J. L., Jamieson, L., Celeste, R. K., Raskin, S. E., Gomaa, N., McGrath, C., & Tiwari, T. (2023). Conceptualizing inequities and oppression in oral health research. Community Dentistry and Oral Epidemiology, 51(1), 28–35. 10.1111/cdoe.12822

Gomaa, N. (2022). Social Epigenomics: Conceptualizations and Considerations for Oral Health. Journal of Dental Research, 101(11), 1299–1306. 10.1177/00220345221110196

Gomaa, N., Nicolau, B., Siddiqi, A., Tenenbaum, H., Glogauer, M., & Quiñonez, C. (2017). How does the social “get under the gums”? The role of socio-economic position in the oral-systemic health link. Canadian Journal of Public Health, 108(3), e224–e228. 10.17269/CJPH.108.5930

Hakeberg, M., & Wide Boman, U. (2017). Self-reported oral and general health in relation to socioeconomic position. BMC Public Health, 18(1), 63. 10.1186/s12889-017-4609-9

Im, J. H. B., Rodrigues, R., Anderson, K. K., Wilk, P., Stranges, S., & Nicholson, K. (2022). Examining the prevalence and correlates of multimorbidity among community-dwelling older adults: Cross-sectional evidence from the Canadian Longitudinal Study on Aging (CLSA) first-follow-up data. Age and Ageing, 51(8), afac165. 10.1093/ageing/afac165

Jin, L., Lamster, I., Greenspan, J., Pitts, N., Scully, C., & Warnakulasuriya, S. (2016). Global burden of oral diseases: Emerging concepts, management and interplay with systemic health. Oral Diseases, 22(7), 609–619. 10.1111/odi.12428

Larvin, H., Kang, J., Aggarwal, V. R., Pavitt, S., & Wu, J. (2021). Multimorbid disease trajectories for people with periodontitis. Journal of Clinical Periodontology, 48(12), 1587–1596. 10.1111/jcpe.13536

Manuel, J. I. (2018). Racial/Ethnic and Gender Disparities in Health Care Use and Access. Health Services Research, 53(3), 1407–1429. 10.1111/1475-6773.12705

McKenna, G., Tsakos, G., Burke, F., & Brocklehurst, P. (2020). Managing an Ageing Population: Challenging Oral Epidemiology. Primary Dental Journal, 9(3), 14–17. 10.1177/2050168420943063

Meyerhoefer, C. D., Pepper, J. V., Manski, R. J., & Moeller, J. F. (2021). Dental Care Use, Edentulism, and Systemic Health among Older Adults. Journal of Dental Research, 100(13), 1468–1474. 10.1177/00220345211019018

Moeller, J., Singhal, S., Al-Dajani, M., Gomaa, N., & Quiñonez, C. (2015). Assessing the relationship between dental appearance and the potential for discrimination in Ontario, Canada. SSM - Population Health, 1, 26–31. 10.1016/j.ssmph.2015.11.001

Nicholson, K., Rodrigues, R., Anderson, K. K., Wilk, P., Guaiana, G., & Stranges, S. (2020). Sleep behaviours and multimorbidity occurrence in middle-aged and older adults: Findings from the Canadian Longitudinal Study on Aging (CLSA). Sleep Medicine, 75, 156–162. 10.1016/j.sleep.2020.07.002

Northridge, M. E., Kumar, A., & Kaur, R. (2020). Disparities in Access to Oral Health Care. Annual Review of Public Health, 41(1), 513–535. 10.1146/annurev-publhealth-040119-094318

Oates, T. W., Guy, V., Ni, K., Ji, C., Saito, H., Shiau, H., Shah, R., Williams, M. A., Blasi, G., & Goloubeva, O. (2023). Meta-regression Analysis of Study Heterogeneity for Systemic Outcomes after Periodontal Therapy. JDR Clinical & Translational Research, 8(1), 6–15. 10.1177/23800844211070467

Ohi, T., Sai, M., Kikuchi, M., Hattori, Y., Tsuboi, A., Hozawa, A., Ohmori-Matsuda, K., Tsuji, I., & Watanabe, M. (2009). Determinants of the Utilization of Dental Services in a Community-Dwelling Elderly Japanese Population. The Tohoku Journal of Experimental Medicine, 218(3), 241–249. 10.1620/tjem.218.241

Peres, M. A., Macpherson, L. M. D., Weyant, R. J., Daly, B., Venturelli, R., Mathur, M. R., Listl, S., Celeste, R. K., Guarnizo-Herreño, C. C., Kearns, C., Benzian, H., Allison, P., & Watt, R. G. (2019). Oral diseases: A global public health challenge. The Lancet, 394(10194), 249–260. 10.1016/S0140-6736(19)31146-8

Polzer, I., Schimmel, M., Müller, F., & Biffar, R. (2010). Edentulism as part of the general health problems of elderly adults*. International Dental Journal, 60(3), 143–155. 10.1922/IDJ_2184Polzer13

Public Health Agency of Canada. (2019, October 31). Canadian Chronic Disease Indicators, 2019 – Updating the data and taking into account mental health [Research]. https://www.canada.ca/en/public-health/services/reports-publications/health-promotion-chronic-disease-prevention-canada-research-policy-practice/vol-39-no-10-2019/chronic-disease-indicators-updating-data-taking-account-mental-health.html

Raina, P., Wolfson, C., Kirkland, S., Griffith, L. E., Balion, C., Cossette, B., Dionne, I., Hofer, S., Hogan, D., van den Heuvel, E. R., Liu-Ambrose, T., Menec, V., Mugford, G., Patterson, C., Payette, H., Richards, B., Shannon, H., Sheets, D., Taler, V., … Young, L. (2019). Cohort Profile: The Canadian Longitudinal Study on Aging (CLSA). International Journal of Epidemiology, 48(6), 1752–1753j. 10.1093/ije/dyz173

Rohani, K., Nicolau, B., Madathil, S., Booij, L., Jafarpour, D., Haricharan, P. B., Feine, J., Alchini, R., Tamimi, F., & de Souza, R. (2023). A Cluster Analysis of Oral and Cognitive Health Indicators: An Exploratory Study on Cholinergic Activity as the Link. JDR Clinical & Translational Research, 23800844231190834. 10.1177/23800844231190834

Rothman, K. J. (2002). Measuring interactions. Epidemiology: An introduction (New York, Oxford University Press).

Sabbah, W., Gomaa, N., & Gireesh, A. (2018). Stress, allostatic load, and periodontal diseases. Periodontology 2000, 78(1), 154–161. 10.1111/prd.12238

Scannapieco, F. A., & Cantos, A. (2016). Oral inflammation and infection, and chronic medical diseases: Implications for the elderly. Periodontology 2000, 72(1), 153–175. 10.1111/prd.12129

Shaw, L. J., Pepine, C. J., Xie, J., Mehta, P. K., Morris, A. A., Dickert, N. W., Ferdinand, K. C., Gulati, M., Reynolds, H., Hayes, S. N., Itchhaporia, D., Mieres, J. H., Ofili, E., Wenger, N. K., & Bairey, M. C. N. (2017). Quality and Equitable Health Care Gaps for Women. Journal of the American College of Cardiology, 70(3), 373–388. 10.1016/j.jacc.2017.05.051

Sheiham, A., & Watt, R. G. (2000). The Common Risk Factor Approach: A rational basis for promoting oral health. Community Dentistry and Oral Epidemiology, 28(6), 399–406. 10.1034/j.1600-0528.2000.028006399.x

StataCopr LLC. (2022). Statistical software for data science—About us. https://www.stata.com/

Statistics Canada. (2023, November 6). The Daily—More than one-third of Canadians reported they had not visited a dental professional in the previous 12 months, CCHS 2022. https://www150.statcan.gc.ca/n1/daily-quotidien/231106/dq231106a-eng.htm

Tsakos, G., & Quiñonez, C. (2013). A sober look at the links between oral and general health. Journal of Epidemiology and Community Health, 67(5), 381–382. 10.1136/jech-2013-202481

Tsakos, G., Watt, R. G., & Guarnizo-Herreño, C. C. (2023). Reflections on oral health inequalities: Theories, pathways and next steps for research priorities. Community Dentistry and Oral Epidemiology, 51(1), 17–27. 10.1111/cdoe.12830

Vandenbroucke, J. P., Elm, E. von, Altman, D. G., Gøtzsche, P. C., Mulrow, C. D., Pocock, S. J., Poole, C., Schlesselman, J. J., & Egger, M. (2007). Strengthening the Reporting of Observational Studies in Epidemiology (STROBE): Explanation and Elaboration. Annals of Internal Medicine, 147(8), W-163. 10.7326/0003-4819-147-8-200710160-00010-w1

Watt, R. G., & Aida, J. (2022). Time to take oral health seriously. The Lancet Healthy Longevity, 3(11), e727–e728. 10.1016/S2666-7568(22)00246-X

Watt, R. G., & Serban, S. (2020). Multimorbidity: A challenge and opportunity for the dental profession. British Dental Journal, 229(5), Article 5. 10.1038/s41415-020-2056-y

Watt, R. G., Venturelli, R., & Daly, B. (2019). Understanding and tackling oral health inequalities in vulnerable adult populations: From the margins to the mainstream. British Dental Journal, 227(1), Article 1. 10.1038/s41415-019-0472-7

WHO. (2022). The Global Status Report on Oral Health 2022. https://www.who.int/team/noncommunicable-diseases/global-status-report-on-oral-health-2022

Zelig, R., Goldstein, S., Touger-Decker, R., Firestone, E., Golden, A., Johnson, Z., Kaseta, A., Sackey, J., Tomesko, J., & Parrott, J. S. (2022). Tooth Loss and Nutritional Status in Older Adults: A Systematic Review and Meta-analysis. JDR Clinical & Translational Research, 7(1), 4–15. 10.1177/2380084420981016

Zhao, D., Zhen, Z., Pelekos, G., Yiu, K. H., & Jin, L. (2019). Periodontal disease increases the risk for onset of systemic comorbidities in dental hospital attendees: An 18-year retrospective cohort study. Journal of Periodontology, 90(3), 225–233. 10.1002/JPER.18-0224

